# Assessing Multimodal AI for Visual Information Extraction of Pharmacology

**DOI:** 10.64898/2026.01.15.26344119

**Authors:** Israel O. Dilan-Pantojas, Phu T. Duong, Kevin Lopes, Richard D. Boyce

**Affiliations:** Department of Biomedical Informatics, University of Pittsburgh, Pittsburgh, PA, USA; School of Mathematics Sciences, Rochester Institute of Technology, Rochester, NY, USA

## Abstract

While Americans are using herbal dietary supplements (natural products) more than ever, the consumption of natural products with prescription drugs can lead to harmful interactions. Pharmacovigilance of natural products depends on careful expert review and interpretation of a wide variety of evidence. In prior work, we demonstrated the value of knowledge graph (NP-KG) for assisting with natural product safety investigations. However, scaling the NP-KG from 33 natural products to the thousands on the market requires computer-assisted data extraction, particularly from visual elements (figures or tables) of pharmacology literature. We evaluated the accuracy and resilience of 8 open- and closed-source multimodal models by performing visual information extraction from select tables and images. The best performing models could accurately extract 90% of tabular data and 45% of data reported figures with a modified relative error rate of 0.05. Image resolution and information density were primary hindrances to better extraction performance.

## Introduction

The use of herbal dietary supplements continues to grow with $12.6 billion in sales in 2023, as reported by HerbalGram in 2024.^1^ Of concern, the consumption of these supplements, often referred to as “natural products”, may potentially cause interactions with prescription medications. The natural product-drug interactions (NPDIs) may lead to serious adverse health effects or even death.^2^ As the market for natural products continues to grow, the importance of NPDI post-market safety/pharmacovigilance increases. Unfortunately, the field is hindered by the limited quality and coverage of data available to study these interactions.^2–4^ Natural products are not regulated like prescription drugs, so high-quality clinical trial data is relatively rare. More available is evidence from *in vitro* screening of natural product complex mixtures and constituents, published clinical case reports, and spontaneously reported safety reports. However, these sources require careful expert review and interpretation to discern cause from association.

Our prior work addressed the highly siloed and fragmented nature of NPDI evidence by developing a Natural Product Knowledge Graph (NP-KG). We demonstrated the application of NP-KG to identify known pharmacokinetic interactions between natural products and pharmaceutical drugs mediated by drug-metabolizing enzymes and transporters.^5^ We also evaluated the ability of several knowledge-graph embedding methods to improve NPDI prediction with NP-KG.^6^ While promising advances, our attempts to scale NP-KG have been hindered by the fact that much of the relevant NPDI evidence in published sources is represented as visual elements such as figures and tables, whose extraction is notoriously difficult to automate. Currently, including data from visual elements into NP-KG requires labor-intensive manual annotation. In this study, we seek to determine whether modern Visual Information Extraction (VIE) methods can help overcome this knowledge maintenance bottleneck, enabling us to scale NP-KG from the 33 natural products in version 3 to all natural products patients use for perceived potential medicinal benefit.

The primary task of a VIE system is the computational extraction of information from visual elements such as diagrams, graphs, figures, and tables.^7–9^ Information may be derived from multiple aspects of these elements. One straightforward example involves extracting textual or numerical data that is explicitly presented in an image. More nuanced examples involve recovering implicitly encoded qualitative or quantitative information from the structure, layout, or visual style of the element, like the row-column organization of tables, the (x,y) coordinate pairs in plots, or the relative heights of bars in bar charts. For this work, the terms charts, plots, and graphs (listed here in order of generality, with chart being the most general) are used interchangeably. In practice, VIE for figures and VIE for tables are often addressed separately, reflecting fundamental differences in how these visual formats encode and structure information. To varying degrees, VIE can be automated using algorithms and techniques such as rule-based digitization, natural language processing, optical character recognition, and more advanced artificial intelligence.

Manual VIE is inherently tedious and error-prone. Scaling manual VIE to large datasets only amplifies these challenges by requiring substantially more time and effort. Annotation accuracy commonly degrades over prolonged annotation sessions due to distractions or fatigue, increasing the likelihood of human error.^10–12^ As a result, automating the process of information extraction from visual elements has been a long-standing goal of computational systems research. Although automated VIE approaches have advanced considerably, they continue to exhibit notable limitations, particularly in extracting information from figures and tables.^13^ Understanding and mitigating these limitations is essential to producing high-quality, reliable information resources for their inclusion in pharmacology research resources like NP-KG.

For decades, digitization has enabled partial automation of VIE. Digitization tools allow users to extract quantitative information from existing graphs by manually identifying reference points and other key features on the original image and then tracing curves or data values. Pixel-distance estimation converts these traced paths into quantitative values relative to the annotated reference points.^14^ More recent digitization software has increased automation through techniques such as color and contrast analysis, which compare background-foreground differences to identify chart components such as groups, series, plots, or quadrants, depending on the figure type.^15,16^ Rule-based processing pipelines allow VIE systems to operate across diverse figure classes (e.g., bar charts, pie charts, maps), though significant limitations remain. A common requirement across digitization systems is manual annotation of key reference or anchor points before automated extraction can proceed. False positives commonly arise during automated annotation, for instance, when legend entries are misinterpreted as data points or when visual noise (e.g., ink artifacts, speckles) is incorrectly captured as part of the plotted data. Modern digitization systems further refine extracted values using machine-learning-based post-processing, such as clustering algorithms that smooth series traces and help reduce false positives.^17–19^

Semi-automated VIE through digitization is particularly useful for visual elements that follow consistent structural and stylistic conventions. However, its scalability remains limited because digitization still demands substantial user involvement, especially when annotators must verify outputs, correct errors, and manage exceptional cases. Performance is further constrained by the quality and heterogeneity of visual elements; digitization tools typically support only a restricted set of figure types, and plots that deviate from these expectations may be partially or entirely unreadable. Moreover, these systems were not designed initially to interpret plots semantically; that is, they cannot autonomously determine which specific values, comparisons, or relationships are relevant to a given research question. As a result, researchers often need to conduct additional manual work to obtain the precise information required.

**Figure 1.**
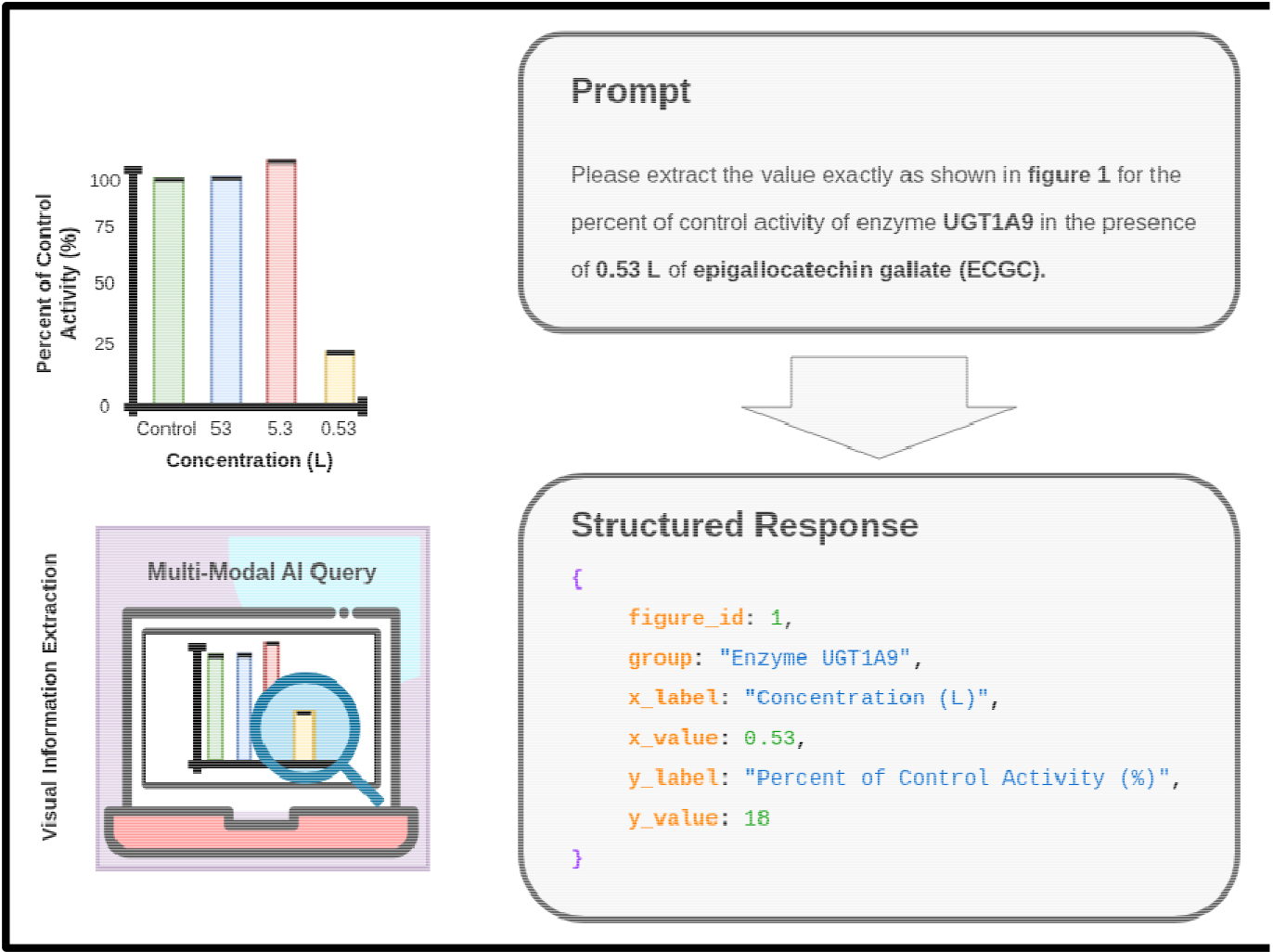
AI-based visual information extraction with structured outputs.

As machine learning (ML) methods continue to advance, more ways to classify and process different visual elements become possible. These methods have expedited automation and enabled the intelligent processing of information from a broader range of visual elements.^16,20^ Among the earliest and most widely used ML approaches for VIE is optical character recognition (OCR), which automatically identifies and extracts alphanumeric characters embedded within an image. However, OCR-based VIE is inherently limited: it cannot recover the non-narrative information encoded through structure, geometry, or layout, such as bar heights in bar charts or relationships among row–column pairs in tables. To compensate for these gaps, OCR is often combined with additional approaches, including natural language processing (NLP). NLP is commonly used to extract information from narrative text,^21^ and can also support VIE when tables are provided in structured formats such as HTML or XML.^22^ Although these techniques help address specific challenges, extracting information from irregular, visually complex, or unstructured tables and figures remains beyond the capabilities of current OCR- and NLP-based systems.

Researchers are increasingly leveraging AI to tackle these challenging visual data representations by incorporating large language models (LLMs) to enhance VIE workflows. LLMs have been successfully used in the pharmacology space to extract pharmacological information from clinical notes involving dietary supplements.^23^ However, this work did not incorporate a VIE component. An early LLM-based system, DePlot^24^, combines OCR with an LLM: OCR first extracted text from table images, which was then submitted to an LLM for question answering. These hybrid approaches improved VIE for tables by exploiting the LLMs’ ability to organize text and infer missing information from their learned representations. However, because they rely primarily on textual inputs, they fail to capture crucial visual information encoded through style, spatial arrangement, or layout.

To overcome this limitation, newer methods integrate convolutional neural networks (CNNs), vision transformers (VTs), or other deep learning architectures with LLMs, enabling models to process both the visual and structural properties of figures and tables. For instance, the DONUT^25^ model improves upon DePlot by eliminating the OCR component and instead using a VT to directly extract visual information into embeddings. These embeddings are passed to an LLM for downstream tasks such as VIE and question answering. Approaches of this kind, broadly known as Vision-Language Models (VLMs) or Multimodal Large Language Models (MLLMs), have significantly advanced the extraction of complex information representations.

Although the specific models described thus far do not yet achieve the performance required for many real-world challenges, transformer-based architectures that jointly encode visual and textual information have yielded notable progress in visual document understanding. These models can capture layout, color, structure, and other stylistic cues within their embeddings, allowing them not only to extract explicit information presented in an image but also to infer implicit relationships described or implied by accompanying text. Consequently, they enable automated VIE across visual elements with high variability, including relative positioning of components, bar heights in shaded plots, point coordinates in scatterplots, and trend patterns within lines or curves.

A key remaining limitation of transformer-based embeddings and multimodal models for VIE is their largely uncharacterized performance under different visual, structural, and contextual conditions. Prior work has evaluated the accuracy of related capabilities, such as question answering and summarization, by generating captions for a reference set of scientific plots.^26^ Although recent models demonstrate promising zero-shot performance on visual document understanding tasks, including document question answering and retrieval, it is unclear how well these models generalize to the demands of VIE.

The goal of this work is to evaluate the performance and resilience of VIE systematically applied to scientific figures and tables containing pharmacological data, with a focus on natural products and their interactions with prescription drugs. We assess multiple commercial and open-source MLLMs to determine their ability to extract and enhance the representation of information from complex, visually diverse data. To do so, we compare model-extracted values against a newly created human-annotated reference set. By improving the accuracy and robustness of machine-based extraction, we aim to enhance the overall quality and reliability of the information captured from these visual elements.

## Methods

To assess the performance and resilience of IE across different AI models, we compare the machine-extracted results with a human-annotated reference set. The extraction experiment is focused on and restricted to two visual element types: figures and tables. To derive information on typical failure modes across the chosen models, we report results from multiple metrics for each element type, and in the discussion, we examine indicators of the elements’ visual complexity, such as visual clarity and information density.

### Creation of the reference set

We leveraged the subset of manually extracted data from figures and tables reported in pharmacology studies entered into repo.napdi.org, a publicly accessible repository of NPDI data. The data comes from the visual elements of 45 published pharmacology studies (Table 1). Multiple images could be sampled from each of these papers, and multiple values were often sampled from each image. Therefore,the reference set contains multiple rows with values from images of tables or from figures corresponding to graphs, plots, or charts. The code and references set are available in Zenodo (10.5281/zenodo.17795454). Eight different types of experimental data are represented in the reference set: *in vitro* enzyme inhibition, induction, and kinetics, *in vitro* transporter inhibition, induction, and kinetics, *in vivo* enzyme kinetics, and *in vivo* human clinical interaction studies (Table 2). The selected sample represents a wide range of styles, layouts, and structures for both figures and tables. During the annotation process, the annotators measured the percent agreement between their original annotation values. The values on which annotators disagreed were adjudicated by either reaching an agreement between annotators or taking the average value between the suggested annotations.

**Table 1.** Reference set images breakdown.

|  | Counts |
| --- | --- |
| Distinct Images of Figures | 43 |
| Distinct Images of Tables | 40 |
| Total Images | 83 |

**Table 2.** Breakdown of queries by experiment type.

| Experiment type | Figure | Table | Total |
| --- | --- | --- | --- |
| *in vitro* enzyme inhibition | 113 | 167 | **280** |
| *in vitro* enzyme induction | 2 | 0 | **2** |
| *in vitro* transporter inhibition | 11 | 11 | **22** |
| *in vitro* transporter kinetics | 8 | 9 | **17** |
| *in vivo* interaction | 14 | 123 | **137** |
| *in vivo* pharmacokinetics | 0 | 39 | **39** |
| Total | **162** | **371** | **533** |

Our goal was to evaluate data extraction performance and not tasks such as automatically identifying relevant visual elements. So, each model shown in Table 3 was prompted to extract data explicitly present in the visual elements. We reviewed each model’s output against the reference set values. We used the Pydantic AI v1.25 Python module for prompt execution and processing of the structured JSON output requested from each model.

**Table 3.** Models included in the evaluation.

| Inference Provider | Model Company | Model Name | Context Window | Number of Parameters |
| --- | --- | --- | --- | --- |
| AWS Bedrock | Anthropic | Claude Sonnet 3.7 | 128K | * |
|  |  | Claude Sonnet 4.0 | 1M | * |
|  | AWS | Nova Pro | 300K | * |
|  |  | Nova Premier | 1M | * |
|  | Meta | Llama 3.2 | 128K | 90B |
|  |  | Llama 4 Scout | 10M | 109B |
|  |  | Llama 4 Maverick | 1M | 400B |
| Open AI API | Open AI | GPT-4o | 128K | * |
|  |  | GPT-5 | 400K | * |
| Google Vertex | Google | Gemini 2.5 Pro | 1M | * |

*\*The actual number of parameters for this model has not been made publicly available.*

### VIE pipeline

The annotation process yielded a reference set table, with each row corresponding to a specific value manually extracted from a visual element. To generate inference results, we iterated each row in the reference set, querying the visual elements from each source. To create the queries, prompt templates corresponding to the experiment type of the row were filled with the information from the row, such that the information extraction task consisted of extracting the necessary information for (x,y) or (row, column) pairings, formatted as (x,?), or a (row, ?), respectively. Sufficient information was provided such that the x and y axes or row/column headers were unambiguously specified in the prompt. Models were instructed to extract only the specific value corresponding to “?” in the (x, “?”) or (row, “?”) pairings, and to provide a structured output compliant with a JSON schema for figures or tables, depending on the element type.

### Metrics

To evaluate the accuracy of the information extracted by the models, we compare the relative difference between the extracted values 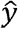 and the target value *y* from the reference set. Here we report both absolute error (Equation 1) and a modified relative error (MRE, Equation 2), used to better quantify the extraction error relative to the magnitude of *y*. The decision to use continuous error metrics is relevant for evaluating extraction from figures because there is some degree of subjective interpretation of the values. The relative error metric was modified with a piecewise definition that allows the evaluation of *y* = 0 was defined. Additionally, we report tolerance-based accuracy between 0.0 and 0.25 MRE to provide a binary classification of the results.

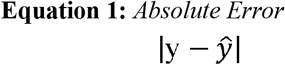

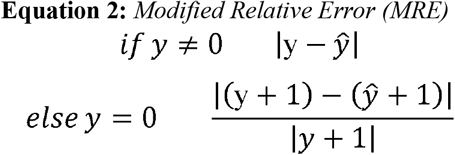

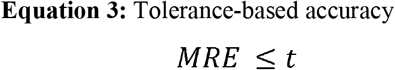

## Results

The percentage agreement between the human annotators on the reference set for the figures was 69% and for the tables it was 100%. It is important to note that the difference in manually annotated values was for the most part within ±2 the target value. So, disagreement for figures represents the range for subjective interpretation of where values land relative to axis ticks on a figure. So, disagreement for figures represents the range for subjective interpretation of where values land relative to axis ticks on a figure. A similar pattern was also observed in most central tendency statistics from VIE experiments (Tables 4 and 5), where models show smaller errors in the tables than in the figures. Except for Llama 4 Maverick, which produced valid outputs for only 515 queries (failed to produce outputs for 18 figure queries), all other models produced valid structured outputs for 533 queries.

**Table 4.** Absolute error from visual information extraction the model failed to produce valid structured outputs for 18 queries.

| Model | Absolute Error |  |  |  |  |  |  |  |  |  |
| --- | --- | --- | --- | --- | --- | --- | --- | --- | --- | --- |
|  | Tables |  |  |  |  | Figures |  |  |  |  |
|  | Mean | Median | Std Dev | Max | Min | Mean | Median | Std Dev | Max | Min |
| Claude Sonnet 3.7 | 9.58 | 0.00 | 78.180 | 1397.60 | 0.00 | 32.35 | 2.00 | 294.46 | 3750.00 | 0.00 |
| Claude Sonnet 4.0 | 83.89 | 0.00 | 1171.40 | 22403.00 | 0.00 | **18.53** | 3.00 | **68.57** | **750.00** | **0.00** |
| Nova Pro | 21.58 | 0.00 | 225.36 | 4213.00 | 0.00 | 52.55 | 14.50 | 276.85 | 3250.00 | 0.00 |
| Nova Premier | **4.75** | **0.00** | **22.29** | **199.98** | **0.00** | 36.36 | 5.00 | 227.80 | 2750.00 | 0.00 |
| Llama 3.2 | 137.49 | 0.00 | 1673.46 | 23196.00 | 0.00 | 80.71 | 16.50 | 611.07 | 7750.00 | 0.00 |
| Scout | 147.78 | 0.00 | 1703.91 | 23222.00 | 0.00 | 52.18 | 10.00 | 413.12 | 5250.00 | 0.00 |
| Maverick * | 8.01 | 0.00 | 54.63 | 969.21 | 0.00 | 61.59* | 10.00* | 448.16* | 5250.00* | 0.00* |
| GPT-4o | 15.71 | 0.00 | 82.26 | 993.68 | 0.00 | 42.66 | 14.00 | 235.74 | 2750.00 | 0.00 |
| GPT-5 | 8.83 | 0.00 | 55.44 | 978.60 | 0.00 | 20.65 | 8.00 | 98.96 | 1250.00 | 0.00 |
| Gemini 2.5 Pro | 7.20 | 0.00 | 54.91 | 978.60 | 0.00 | 22.29 | **2.00** | 215.94 | 2750.00 | 0.00 |

**Table 5.** Modified relative error from visual information extraction.

| Model | Modified Relative Error |  |  |  |  |  |  |  |  |  |
| --- | --- | --- | --- | --- | --- | --- | --- | --- | --- | --- |
|  | Tables |  |  |  |  | Figures |  |  |  |  |
|  | Mean | Median | Std Dev | Max | Min | Mean | Median | Std Dev | Max | Min |
| Claude Sonnet 3.7 | 0.12 | 0.00 | 0.71 | 9.11 | 0.00 | 0.53 | 0.07 | 2.13 | 25.00 | 0.00 |
| Claude Sonnet 4.0 | 26.08 | 0.00 | 339.95 | 6078.85 | 0.00 | 1.17 | 0.09 | 6.73 | 75.00 | 0.00 |
| Nova Pro | 1.37 | 0.00 | 22.75 | 438.13 | 0.00 | 2.78 | 0.36 | 10.73 | 90.00 | 0.00 |
| Nova Premier | 0.12 | 0.00 | **0.60** | 9.00 | 0.00 | 1.03 | 0.18 | 6.01 | 75.00 | 0.00 |
| Llama 3.2 | 0.59 | 0.00 | 4.08 | 53.09 | 0.00 | 1.80 | 0.45 | 4.11 | 26.41 | 0.00 |
| Scout | 0.49 | 0.00 | 3.95 | 55.03 | 0.00 | 1.01 | 0.33 | 2.44 | 20.00 | 0.00 |
| Maveric* | 0.16 | 0.00 | 1.26 | 23.48 | 0.00 | 1.53* | 0.26* | 7.15* | 80.00* | 0.00* |
| GPT-4o | 0.20 | 0.00 | 0.69 | 8.24 | 0.00 | 1.06 | 0.36 | 2.19 | 20.00 | 0.00 |
| GPT-5 | 0.13 | 0.00 | 0.54 | 9.00 | 0.00 | 0.84 | 0.20 | 5.51 | 70.00 | 0.00 |
| Gemini 2.5 Pro | **0.11** | **0.00** | 0.68 | **9.00** | **0.00** | **0.34** | **0.05** | **0.96** | **6.69** | **0.00** |

For Tables 4 and 5, we have noted the best score for each statistic in bold, and the overall best-performing model for the metric has been highlighted in bold. The absolute error values resulting from the VIE extraction experiments for tables show that the AWS Nova Premier model achieved the lowest mean absolute error and standard deviation values (Table 4). All models achieved minimum and median absolute errors of 0.0 in table extraction. Models Claude Sonnet 3.7, Llama 4 Maverick, GPT-5, and Gemini 2.5 Pro also performed relatively well. For the absolute error on VIE, based on the figures, Claude Sonnet 4.0 performed best overall. For figures, all models only achieved a minimum absolute error of 0.0. Here, models GPT-5 and Gemini 2.5 Pro also performed relatively well. We should note that the results for extraction from figures by Llama 4 Maverick are marked with an asterisk (*) since

The MRE values from the VIE extraction experiments for the tables show that, when we consider the magnitude of the error relative to the target value, Gemini 2.5 Pro produced the least overall error delta (Table 5). The AWS Nova Premier model achieved the lowest standard deviation values and also produced low error values. Interestingly, when considering the relative magnitude of the extraction errors from tables, Claude Sonnet 4.0 performed substantially worse than all other models on VIE from tables. Gemini 2.5 Pro also produced the least overall error in the extraction from figures. Models Claude Sonnet 3.7 and GPT-5 also performed relatively well.

In Figure 2, the mean values are shown as horizontal black bars. Since the median was zero across all models, they are not shown in the plot. The overall distribution of extraction deltas from tables across all model groups is closer to 1.0. Claude Sonnet 3.5, AWS Nova Premier, Llama 4 Maverick, GPT-4, GPT-5, and Gemini 2.5 Pro all have similar mean values. The distribution for Gemini 2.5 Pro, which had the smallest relative error, shows that most of its relative errors are smaller in magnitude than the target value.

**Figure 2.**
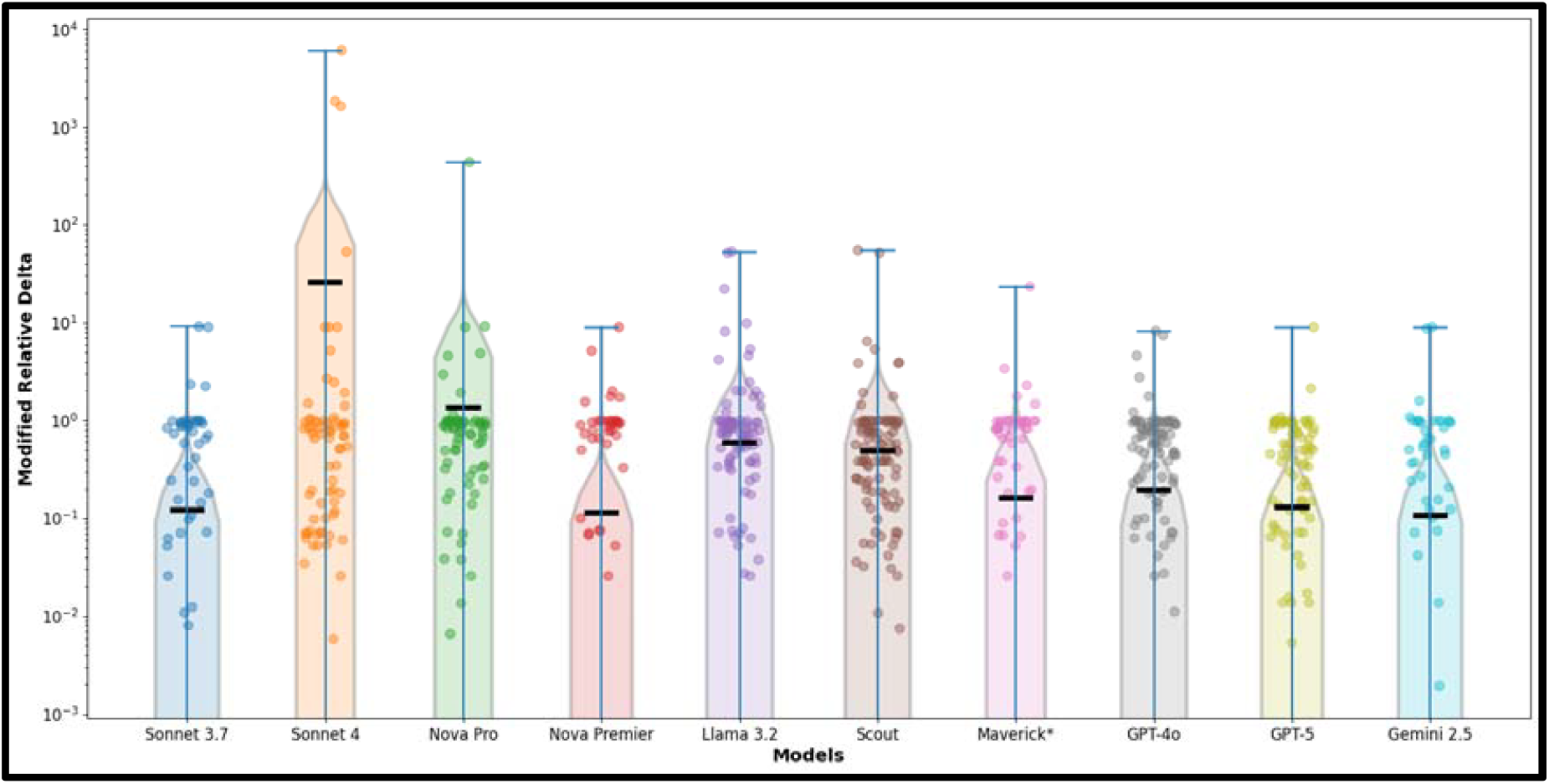
Comparison of models’ extraction errors on tables.

In Figure 3, the means are also shown as horizontal black bars, and medians are shown as horizontal blue bars. The overall distribution of deltas for extraction from figures across all models ranged from 10□^2^ to 10□. The Sonnet 3. and Gemini 2.5 Pro models have the lowest mean and median errors. Gemini 2.5 Pro also seems to have performed best on VIE form figures.

**Figure 3.**
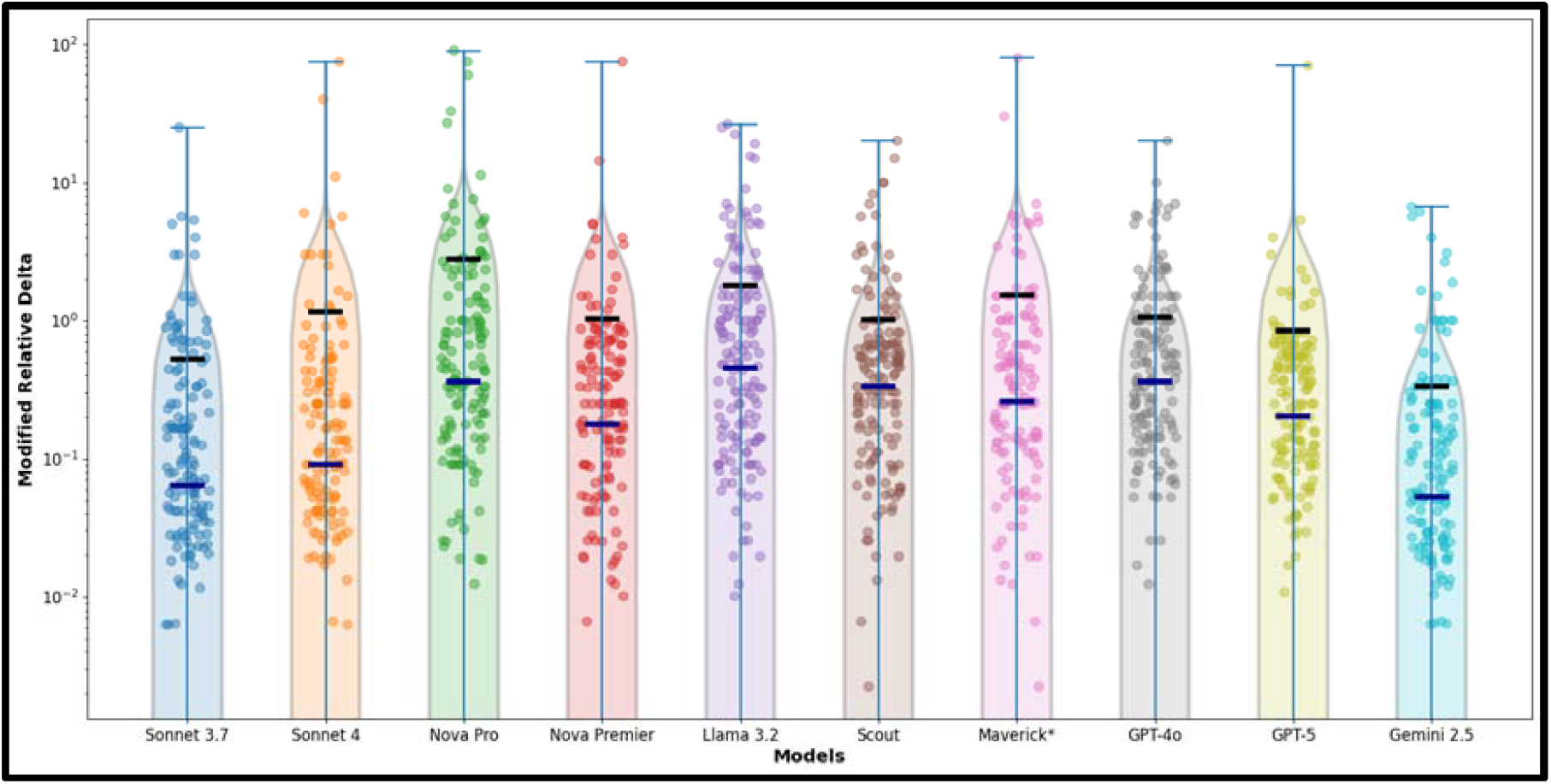
Comparison of models’ extraction errors on figures.

To simplify the practical interpretation of the continuous error in the MRE metric, Table 6 presents a tolerance-based categorical binarization across multiple thresholds, reported as tolerance-based accuracy. Values with an MRE smaller than the threshold are considered correct or acceptable extractions. Looking at the extraction from tables, both Gemini 2.5 Pro and AWS Nova Premier performed very similarly across all reported thresholds, achieving 90% accuracy when the threshold is set to 0.05 times the target magnitude. For figures, Gemini 2.5 Pro also achieves the highest accuracy, correctly extracting 58% of the queries with a threshold of 0.10 times the target’s magnitude.

**Table 6.**
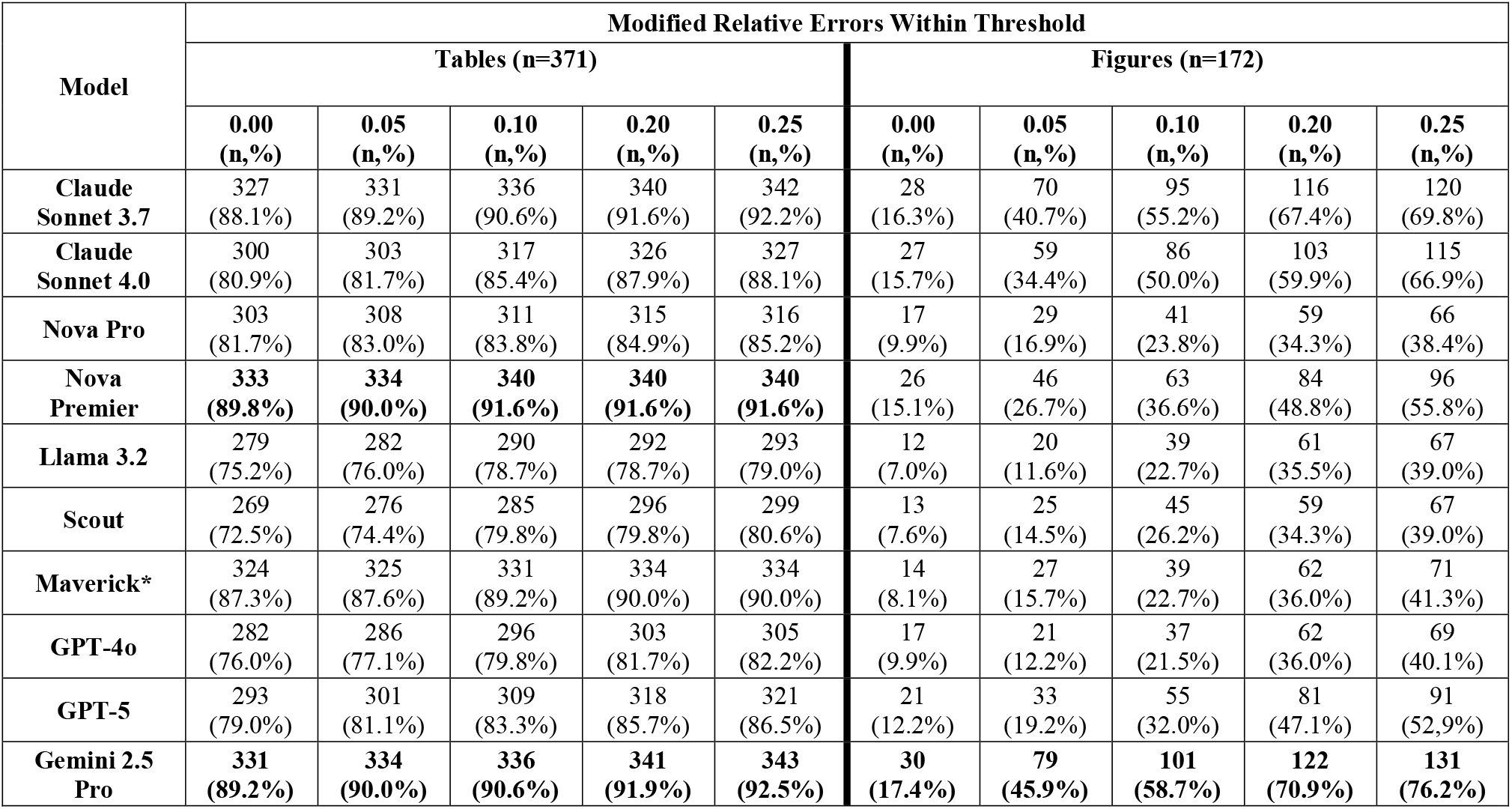
Tolerance-based accuracy with variable thresholds.

## Discussion

Our experiments demonstrate that VLMs and MLLMs can perform VIE across a wide range of scientific figures and tables, with varying degrees of accuracy. Most models achieved a mean MRE smaller than 0.5 times the magnitude of the target value, both for figures and tables. The best performing models could accurately extract 90% of tabular data and 45% of data reported figures with a modified relative error rate of 0.05. This is very result suggests that nearly fully automated table VIE is within reach. However, the values extracted from figures are notoriously difficult to extract precisely, as evidenced by the higher mean MRE when compared to extraction from tables. These findings suggest that, for our use case, we will need to develop a semi-automated workflow for figure data extraction.

The primary source of deviation in extraction accuracy was visual clarity, particularly the image size and resolution. For images with low visual clarity, models often extracted incorrect values from tables, confusing “Ki” with “Km” or numbers like “1399” with “1599”. The secondary source was the way information was conveyed in the plots. For example, zero-valued bars in barplots are deduced by the absence of a bar, or how the information for the identity of a bar or series might only be described with text in the figure description. Another element with a significant impact on extraction accuracy was the information density of the plot, specifically how many series or bars are closely grouped, similarly named, and possibly overlapping. Another factor that seems to have a significant impact on extraction accuracy is the information density of the plot, specifically how many series or bars are closely grouped, similarly named, or overlap. Another significant source of deviation between target and inferred values was the difference in nomenclature across table headers or plot axes. In general, closed-sourced models adhered to the requested output structure reliably. The open-sourced models generated inconsistent structured output that required substantially more processing and error handling.

Due to the lack of control over API service bandwidth throttling and variable network latency, the runtimes of each experiment are not reported. Most models processed the entire dataset within 45 minutes, with notable exceptions: GPT-5 and Gemini 2.5 Pro, which experienced severe throttling and took several hours to complete. Nonetheless, this is considerably less time than the weeks it took to put together the reference set.

During testing, we identified a secondary use of the extraction pipeline: extractions with a large MRE sometimes indicated annotation errors in the reference set. In essence, AI-powered VIE not only serves as an information extraction methodology but could also serve as a partial step in a data validation pipeline. Thus, improving the quality of data being entered into an information resource, such as NP-KG.

In this paper, we did not compare the cost of extraction with multi-modal AI models to that of AI-based VIE approaches, such as digitization and OCR, or other systems, such as AWS Bedrock’s Intelligent Document Processing. We believe these approaches are robust but rigid, meaning that as the variance between images increases, the reliability of the existing automation with these systems decreases. This limitation is not very significant when processing large datasets of identical or similar images. In many cases, it might be possible to adapt the extraction process to newer variations by spending more time developing additional pipelines. Another limitation of this study is that we did not extensively test the models’ ability to identify synonyms or extract related content concepts; in fact, we explicitly aimed to avoid this label-transformation task and instead opted for more precise, accurate labeling of the table headers and figure axes in the training set. Notice that this label matching can be easily achieved by executing multiple calls to the model for that specific task of mapping labels. Alternatively, a label-matching agent could map a general label to the particular table headers or figure axes labels in the image. We also did not extensively explore the role of color, simply because we did not have sufficient plots with different-colored elements in the original data set. However, we noticed that images with monochrome shaded bars in the barplot were often among the larger VIE errors.

## Conclusion

The results from these experiments show the capability of MLLMs to perform relatively accurate VIE from images of scientific figures and tables. Both Gemini 2.5 Pro and AWS Nova Premier performed very similarly in table extraction accuracy. While Gemini 2.5 Pro outperformed the other models in extraction accuracy for figures. In addition, we list the factors we encountered that had a high potential to affect the extraction’s accuracy, namely visual clarity and information density. Inadvertently, the current prototype extraction system proved helpful as a partial validation tool during the annotation of the reference set. Similar systems can help improve the quality and throughput of data creation for systems such as NP-KG. The lessons learned from this work will be applied in the creation of future VIE pipelines. Future work will include a structured evaluation that stratifies visual complexity elements. Additionally, we plan to conduct further experiments that leverage Agentic AI pipelines to automate complementary processing steps for more nuanced extraction of the different visual element types.

## Data Availability

All data produced are available online at

https://zenodo.org/records/17795454

## Acknowledgements

Funding from NIH T15 Grants T15LM007059, U54 AT008909, and the University of Pittsburgh School of Pharmacy for providing funding that enabled us to complete this work. Additionally, we would like to thank the AWS Generative AI Innovation Center (GAIIC) for providing advice on the set up and use of the AWS Bedrock Inference API.

